# Metallo-β-lactamase mediated rapid increase in carbapenem resistance of *Pseudomonas aeruginosa*

**DOI:** 10.1101/2024.03.21.24304089

**Authors:** Hasnain Anjum, Shomaia Yasmin Mitu, Md. Shamsul Arefin, Meftahul Jannat Mitu, Md. Sabir Hossain, Salequl Islam, M.A. Karim Rumi, M. Hasibur Rahman

## Abstract

Antibiotic-resistant *Pseudomonas aeruginosa* is a common nosocomial pathogen all over the world. We detected the presence of *P. aeruginosa* in 22% (53 out of 238) of the test samples collected from patients with infections including secondary wound infections, abscesses and urinary tract infections admitted to two academic hospitals in Bangladesh. Resistance to carbapenems (imipenem, and meropenem) was present among 30% (16 out of 53) of these clinical *P. aeruginosa* isolates, which is more than 2-fold higher compared to that of previous studies. Such a rapid increase in carbapenem resistance was mediated by metallo-β-lactamase (MBL). Expression of MBL was detected in 90% (14 out of 16) of these resistant isolates. Molecular analyses revealed that the carbapenem-resistant isolates carried at least one of the MBL variants, either *bla-*VIM or *bla*-NDM-1. All the *bla-*NDM-1 positives carried a 0.5 MDa plasmid. ERIC-PCR revealed the highly heterogeneous nature of the *P. aeruginosa* isolates indicating multiple sources of infection within the hospital. However, the majority of XDR isolates belonged to a single cluster of drug-resistant bacterial infections. These findings indicate that Metallo-β-lactamase (MBL) mediated resistance to carbapenem in *P. aeruginosa* poses a serious threat to the spread of infections among hospitalized patients.

## 1. Introduction

*Pseudomonas aeruginosa* (*P. aeruginosa*) is one of the most common pathogens associated with nosocomial infections (1). This Gram-negative bacterium is a member of the infamous ESKAPE group of bacteria, known for their rapidly increasing antibiotic resistance in healthcare facilities (2, 3). The global priority list of emerging antibiotic-resistant bacteria recognizes *P. aeruginosa* as a critical pathogen that requires an in-depth study(4). It is commonly associated with wound infections, urinary tract infections (UTIs) (particularly catheter-associated UTIs), ventilator-associated respiratory tract infections (RTI), and cystic fibrosis (1, 5).

The carbapenem group of antibiotics (imipenem, meropenem, ertrapenem, and doripenem) is clinically effective against *P. aeruginosa* (5, 6). However, a steep rise in carbapenem resistance has been reported over the last few years (7). Susceptibility to carbapenems is dependent on their entry to bacteria through the outer membrane porin protein OprD (8). Thus, a reduced expression of OprD can lead to carbapenem resistance in *P. aeruginosa* (8). Other mechanisms of carbapenem resistance include decreased permeability of outer membrane porins, increased production of molecular efflux pumps, acquisition of chromosomal cephalosporinases, and expression of broad-spectrum beta-lactamases, especially carbapenemases like metallo-β-lactamases (MBLs)(9). Acquisition of MBLs through horizontal gene transfer has been increasingly identified in Gram-negative isolates from hospitalized patients(10). These enzymes can hydrolyze most of the β-lactams and are not repressed by serine β-lactamase repressors like tazobactam and clavulanates (11). As MBLs depend on bivalent zinc ions for the hydrolysis of β-lactam antibiotics, they are inactivated by metal chelators like EDTA(12).

The origins of MBLs remain unclear; it is most likely transferred from environmental bacteria like *Enterobacteriaceae* (13). The genes encoding MBLs are mainly spread through mobile genetic elements like integrons residing on plasmids or bacterial genomic DNA (13, 14). The majority of the MBLs belong to subclass B1 (15, 16) and the 3 most common MBLs, namely IMP (imipenemase), VIM (Verona integron-encoded metallo-β-lactamase), and NDM(New Delhi metallo-β-lactamase) are well-known for their epidemiological and clinical importance (12, 13, 17). The *bla*-VIM was first detected in *P. aeruginosa* in European countries during the late 1990s, and more than 20 different VIM allotypes have been identified worldwide (13, 18). However, *bla*-NDM-1, which was detected in *K. pneumoniae* in 2008, has been widely disseminated among *Enterobacteriaceae* in India (14). A significant increase in the prevalence of these MBLs has been seen throughout the world, including the United Kingdom and Southeast-Asian countries like India, Pakistan, and Bangladesh (14, 19, 20).

In this study, we investigated the presence of drug-resistant *P. aeruginosa* infection among hospitalized patients in Dhaka, Bangladesh. We observed that resistance to carbapenems has doubled in the past decade (6). Such a rapid increase in carbapenem resistance was mediated by metallo-β-lactamase (MBL). We detected the expression of MBL-variants in carbapenem resistant *P. aeruginosa* isolates and determined the heterogenic nature of the clinical isolates.

## 2. Materials and Methods

### 2.1 Isolation and Characterization of clinical isolates

Clinical samples were collected from patients admitted to two academic hospitals, Enam Medical College Hospital (EMCH) and Gonoshasthaya Samaj Vittik Medical College Hospital (GMCH) located at Savar, Bangladesh. The presence of *P. aeruginosa* was studied in 238 samples consisting of 115 midstream urine, 69 pus, 21 secondary wound infection swab, 12 urinary catheter swab, 10 burn wounds, 9 blood, and 2 tracheal aspirate specimens. *Pseudomonas* Cetrimide Agar (Scharlab S. L., Spain) was used to determine the colony morphology, fluorescence, and pigment production, followed by conventional biochemical tests according to Bargey’s Manual of Systemic Bacteriology for presumptive identification of *P. aeruginosa* as described previously (41, 42). Selected isolates were stored in glycerol broth at −80 °C.

### 2.2 Molecular Identification

Molecular identification was carried out by sequencing 16S rDNA of fourteen representative isolates. DNA templates prepared from the selected isolates were subjected to polymerase chain reaction (PCR) (43). The PCR products were purified using Promega Wizard SV Gel and PCR Clean-up System, USA and sequenced. The identity of the isolates was confirmed by BLAST analysis.

### 2.3. Screening of Carbapenem Resistance

To determine the susceptibility of clinical *P. aeruginosa* isolates to carbapenem antibiotics, Kirby Bauer disc diffusion test was used (44), followed by determination of MIC against Imipenem (IMP 10 µg) and Meropenem (MPM 10 µg). Antibiotic discs were obtained from Oxoid, UK. *P. aeruginosa* PAO1 was used as control. Results were interpreted according to CLSI standards and guidelines [24]. MDR nature of isolates was observed against nine additional antibiotics of six different groups (data not included in this study) and interpreted accordingly (45, 46).

### 2.4. Phenotypic Detection of MBL Production

To detect the production of MBL, imipenem-resistant isolates were subjected to IMP-EDTA double disc synergistic test. The pure cultures of selected isolates were plated on Mueller-Hinton agar (Oxoid, UK) to prepare bacterial lawn. Two imipenem (IMP 10 µg) disks were placed on prepared lawn by at least 20 mm distance. One of the disks was impregnated with 0.5M EDTA to achieve 750 µg/disk concentrations, which can act on MBL by removing zinc ions from the active site of the enzyme, effectively nullifying its activity. The plates were incubated overnight at 37 °C, and zone diameters for both IMP and IMP-EDTA disks were measured. Isolates exhibiting ≥17 mm inhibition zones with IMP-EDTA disc were considered MBL-positive, while isolates with ≤14 mm inhibition zones were considered MBL-negative (47).

### 2.5. PCR Detection of MBL

The presence of genes encoding four different variants of MBLs *bla*-NDM-1, *bla*-SPM, *bla*-IMP, and *bla*-VIM were investigated in all isolates exhibiting phenotypic resistance against imipenem. PCR reagents were obtained from Promega, USA and the primers were designed according to previous publications (24, 48). The primer as well as annealing temperatures are listed in **Table 1**. For each PCR reaction, 2 µl of prepared bacterial DNA was added to 16 µl of prepared PCR-ready mix containing (1X PCR buffer, 2.5mM MgCl2, 0.25 mM dNTP, 1IU of Taq DNA polymerase, 10 pmol of each primer), and deionized water was added to obtain a final volume of 25 µl. The PCR reactions were carried out as indicated in **Table 3**. The PCR products were visualized using a UV transilluminator after electrophoresis in 1% agarose gel.

**Table 1.**
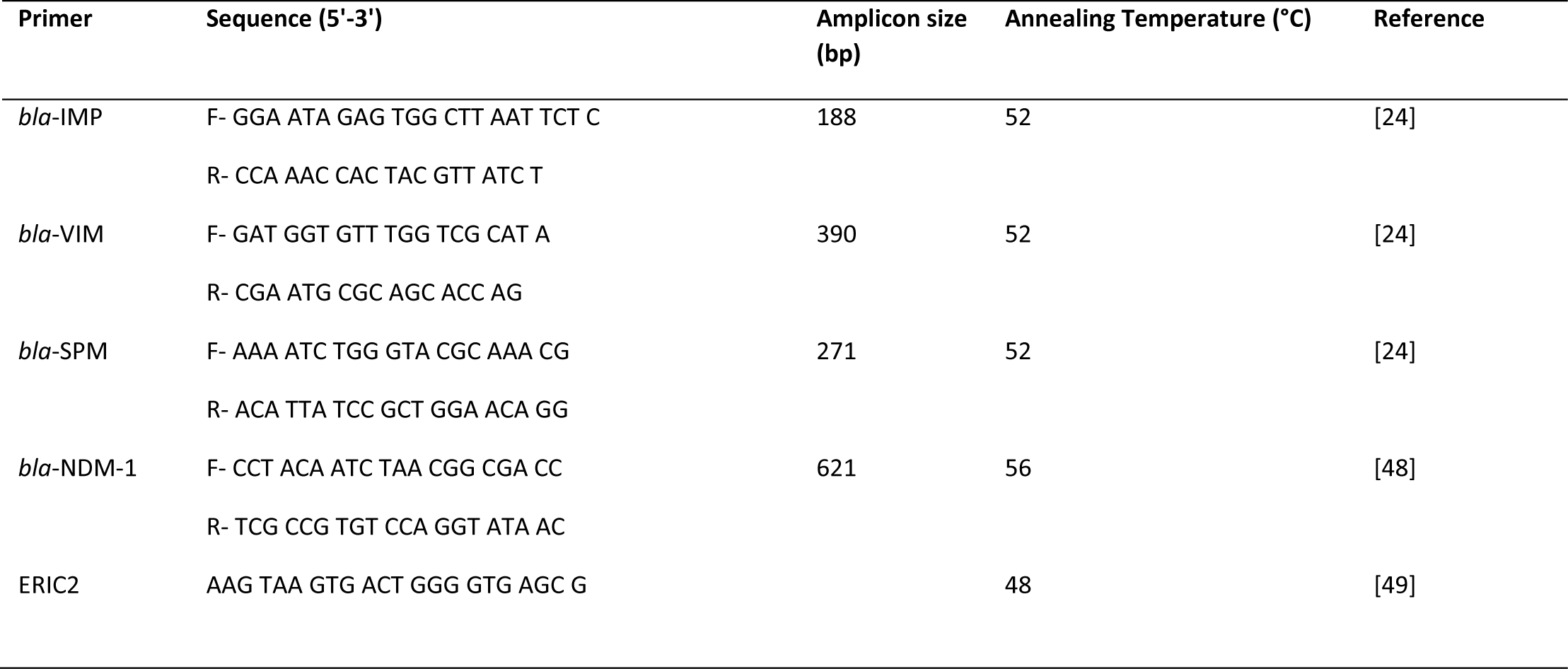
List of PCR primers used in this study.

### 2.6. ERIC-PCR for Species Diversity Analysis

To observe the diversity among the *P. aeruginosa* isolates, ERIC-PCR (Enterobacterial Repetitive Intergenic Consensus-Polymerase Chain Reaction) was performed using the primer sequence mentioned elsewhere (49). PCR products were separated on 2% agarose gel and visualized under a UV transilluminator. The cluster analysis was conducted by using DendroUPGMA (21, 50).

### 2.7. Plasmid Extraction

Plasmids from carbapenem-resistant *P. aeruginosa* isolates were extracted according to the modified hot alkaline lysis method by Kado and Liu (51). Extracted plasmids were electrophoresed in 0.7% agarose gel with ethidium bromide (0.5 mg/mL) and visualized under a UV transilluminator. Reference strains of *E. coli* K-12 V517 and PDK9 were used as control strains for the extraction method and plasmid size marker (52, 53).

### 2.8. Statistical Analysis

A validated questionnaire was used for data collection. Collected data were verified and analyzed by using IBM SPSS Statistics Data Editor (Version 21) and STATA 15.

## 3. Results

### 3.1. Isolation and identification of P. aeruginosa

A total of 238 clinical samples were collected from the patients admitted into two academic hospitals in Savar City, at the outskirts of Dhaka, the capital of Bangladesh. Most of clinical samples were collected from patients with UTI, followed by secondary wound infections and abscesses (**Table 1**). The samples were tested for the presence of *P. aeruginosa* and 53 (∼22%) showed growth on selective cetrimide agar, which were further characterized through standard biochemical tests. Identification of the clinical isolates was confirmed by PCR-based 16SrDNA sequence analyses, which showed a 95 to 99% identity to *P. aeruginosa* sequence (e-value ≤0). We detected that wound infections were more frequently positive for *P. aeruginosa* compared to other infections.

### 3.2. Carbapenem resistance in clinical P. aeruginosa isolates

All 53 isolates of *P. aeruginosa* were tested for susceptibility to carbapenem (imipenem and meropenem) using a disk-diffusion method followed by minimum inhibitory concentration (MIC) assays. Out of 53 isolates, 16 (30%) were resistant to both imipenem and meropenem. However, the antibiotic tolerance levels varied widely in the MIC assay. While 50% (8/16) of carbapenem-resistant isolates showed a moderate resistance (8 µg/ml), the other 50% isolates had a much higher resistance level (≥16 to 128 µg/ml). Among the 16 carbapenem*-*resistant *P. aeruginosa* isolates, 6 were from GMCH (38%), while the rest 10 (62%) were from the EMCH (**Table 1**). Antibiogram profile identified 12 isolates as multidrug resistant (MDR), and another 2 isolates as extensively drug resistant (XDR). Moreover, uniform resistance against third generation cephalosporins like ceftriaxone and cefotaxime and significantly lower susceptibility to important antibiotics like azithromycin and tetracycline was observed, which is alarmingly higher than previous regional reports.

### 3.3. Detection of MBL in P. aeruginosa isolates

All of the 16 imipenem-resistant *P. aeruginosa* isolates were subjected to enzymatic activity and PCR based genotypic detection of MBL variants. Of the 16 isolates, 14 (∼88%) were positive for MBL activity in an imipenem-EDTA double-disk synergistic test. PCR-based detection of the MBL variants, *bla*-VIM, *bla*-NDM1, *bla*-IMP, and *bla*-SPM, showed that 10 out of the 16 (∼63%)resistant isolates carried at least one. While *bla-*VIM was detected in 7 out of 16 isolates (44%), *bla*-NDM-1 was detected in 3 isolates (19.75%) (**Figure 1**). We did not detect any of MBL variants, *bla*-VIM, *bla*-NDM1, *bla-* IMP, and *bla*-SPM in other 6 isolates. Moreover, none of the isolates were multiple MBL genes carrier (**Table 3**).

**Figure 1.**
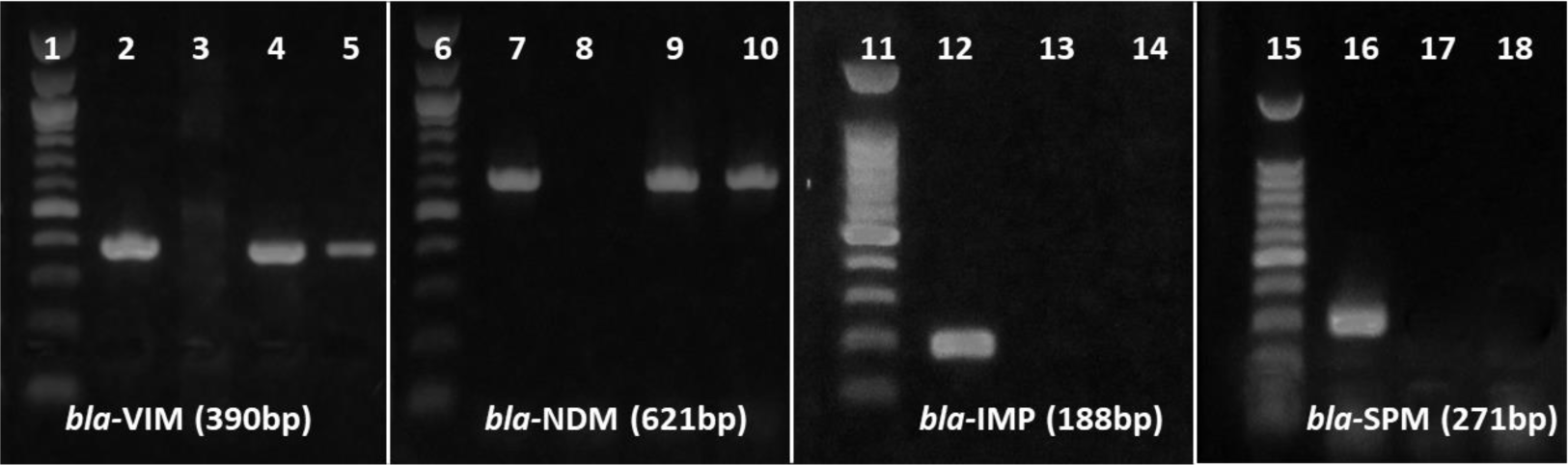
PCR based detection of MBL variants. MBL gene sequences were PCR amplified using the variant**-**specific primers (Table 3). PCR products were electrophoresed through 1% agaroge gels, stained with ethidium bromide, and visualized with UV light. The 100 bp DNA ladder indicates the size of PCR products. Lane 2, 7, 12 and 16 represent variant-specific positive controls, and lane 3, 8, 13 and 17 represent negative controls. No isolates were positive in *bla-*IMP and *bla-*SPM specific PCR.

**Table 2.** Collection of samples and culture results.

| Sample type | Samples collected from EMCH |  | Samples collected from GMCH |  | Total samples | Total *P. aeruginosa* isolates |
| --- | --- | --- | --- | --- | --- | --- |
|  | Sample number | Positive for *P. aeruginosa* | Sample number | Positive for *P. aeruginosa* |  |  |
| Urine | 72 (62%) | 16 (22%) | 43 (38%) | 7 (16%) | 115 | 23 |
| Urinary Catheter | 7 (58%) | 4 (57%) | 5 (42%) | 1 (20%) | 12 | 5 |
| Sec. wound infection | 12 (57%) | 7 (58%) | 9 (43%) | 4 (45%) | 21 | 11 |
| Pus | 37 (54%) | 5 (13%) | 32 (46%) | 5 (15%) | 69 | 10 |
| Burn wound | 8 (80%) | 2 (25%) | 2 (20%) | 0 | 10 | 2 |
| Blood | 9 (100%) | 1 (11%) | 0 | 0 | 9 | 1 |
| Tracheal aspirate | 2 (100%) | 1 (50%) | 0 | 0 | 2 | 1 |
| Total | 147 (62%) | 36 (24%) | 91 (38%) | 17 (19%) | 238 | 53 |

**Table 3.** Antibiotic susceptibility and resistance determinants in Imipenem-resistant clinical *P. aeruginosa*.

| Isolate ID | Specimen (Hospital) | MIC Value (µg/ml) | Phenotypic Detection of MBL Production |  |  | Presence of MBL gene |  |  |  | Plasmid (MDa) |
| --- | --- | --- | --- | --- | --- | --- | --- | --- | --- | --- |
|  |  |  | IMP (Zone, mm) | IMP+ EDTA (Zone, mm) | Interpretation | *bla*-IMP | *bla*-VIM | *bla*-SPM | *bla*-NDM1 |  |
| PA01<sup>a</sup> | Reference | 0.5 | 19 | 20 | Negative | – | – | – | – | – |
| PU1 | Urine (A) | 8 | 0 | 0 | Negative | – | – | – | – | – |
| PU5 | Urine (A) | 8 | 0 | 14 | Negative | – | – | – | – | – |
| PU6 | Urine (A) | 8 | 0 | 15 | Intermediate | – | – | – | – | – |
| PUS1 | Cath. Swb (A) | 8 | 0 | 15 | Intermediate | – | – | – | – | – |
| PS1 | Wound Swb (A) | 16 | 0 | 16 | Intermediate | – | – | – | – | – |
| PS2 | Wound Swb (A) | 8 | 0 | 15 | Intermediate | – | – | – | – | – |
| PU17 | Urine (B) | 128 | 0 | 22 | Positive | – | – | – | + | 0.5 |
| PU26 | Urine (B) | 8 | 0 | 20 | Positive | – | + | – | – | 1.2 |
| PWS12 | Wound Swb (B) | 32 | 0 | 20 | Positive | – | + | – | – | – |
| PWS17 | Cath. Swb (B) | 128 | 0 | 24 | Positive | – | – | – | + | 0.5, 1.2 |
| PWS18 | Cath. Swb (B) | 128 | 0 | 21 | Positive | – | – | – | + | 0.5, 1.2 |
| PWS21 | Wound Swb (B) | 128 | 0 | 24 | Positive | – | + | – | – | 0.2, 1.2 |
| PWS22 | Wound Swb (B) | 16 | 0 | 22 | Positive | – | + | – | – | 0.2, 1.2 |
| PWS23 | Wound Swb (B) | 16 | 0 | 16 | Intermediate | – | + | – | – | – |
| PWS25 | Cath. Swb (B) | 8 | 0 | 16 | Intermediate | – | + | – | – | 0.2, 1.2 |
| PWS26 | Tracheal Asp (B) | 128 | 0 | 23 | Positive | – | + | – | – | – |

### 3.4. Detection of plasmids in MBL positive isolates

Plasmid extraction and gel electrophoresis detected the presence of plasmids in 7 out of the 16 (43%) MBL-positive isolates. Among 7 *bla*-VIM positive isolates, 4 carried a plasmid of 1.2 MDa, while the other 3 isolates did not carry any plamid. In constrast all 3 of the *bla*-NDM1 positive *P. aeruginosa* isolates carried a plasmid of 0.5 MDa (**Table 3**).

### 3.5. ERIC-PCR and cluster analysis

Enterobacterial Repetitive Intergenic Consensus-Polymerase Chain Reaction (ERIC-PCR) was performed to determine the possible epidemiological relationships among the clinical *P. aeruginosa* isolates (21).Our results revealed distinct patterns of DNA, indicating species diversity among carbapenem-resistant *P. aeruginosa* isolates. Highly heterogeneous results indicated that the isolates did not share significant genetic relatedness. Phylogenetic cluster analysis based on unweighted pair group method with arithmetic mean (UPGMA) exhibited the presence of three distinct clusters I-III (**Figure 2**). The majority of the XDR isolates belonged to cluster III, while cluster I and II were composed of MDR isolates.

**Figure 2.**
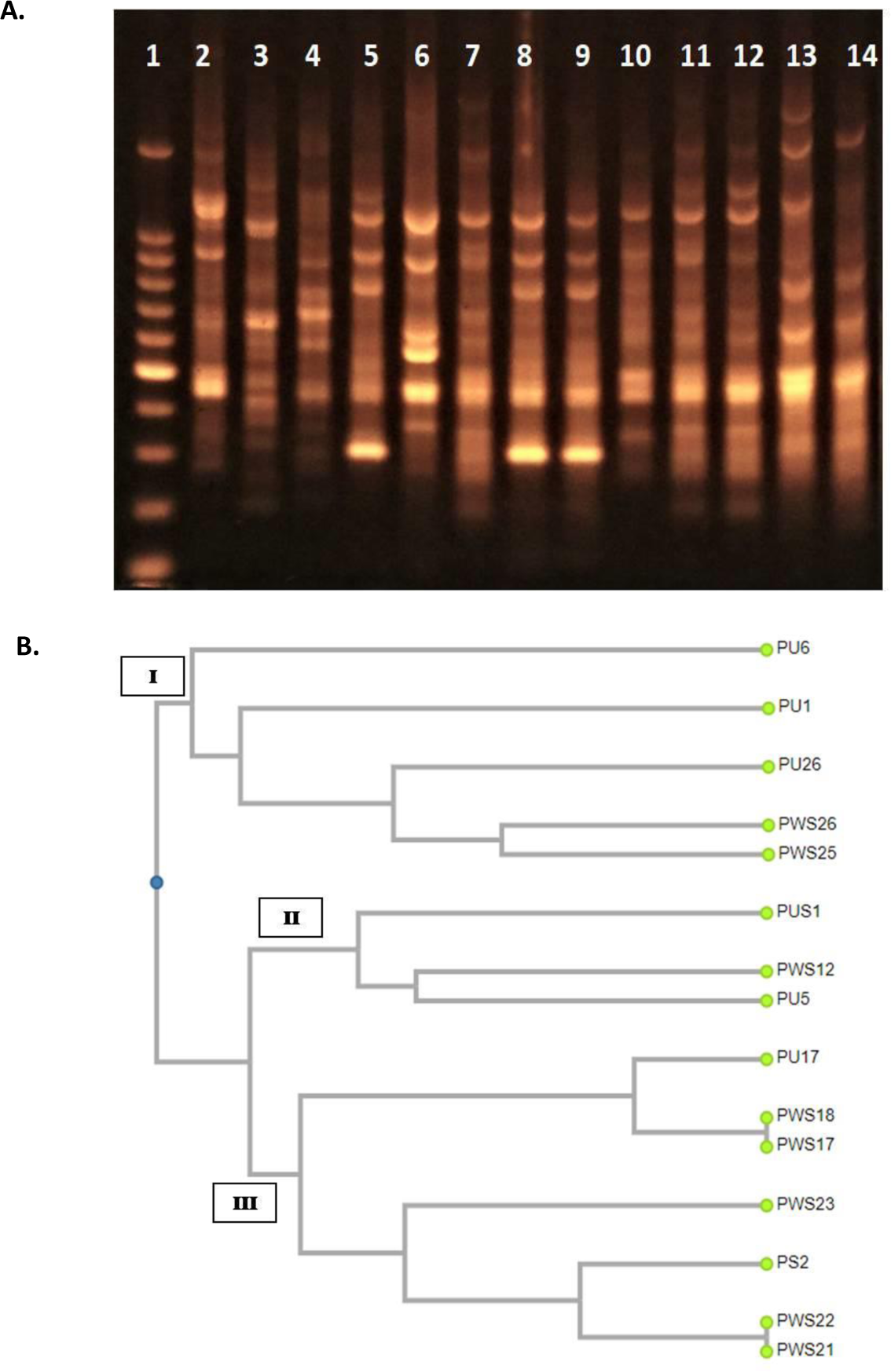
UPGMA cluster analysis of ERIC-PCR results. A). ERIC-PCR products were electrophoresed on 1% agarose gel, stained with ethidium bromide, and visualized on UV light. Lane 1 represent 100 bp DNA ladder; Lane 8 and 9 represent isolates PWS18 and PWS17 respectively showing identical amplification pattern. Similarly, Lane 10 and 11 represent isolates PWS22 and PWS21 respectively showing identical amplification. Rest of the lanes represent unique PCR amplification profiles. B). Dendrogram based on UPGMA cluster analysis of ERIC-PCR results; the names of the analyzed strains are mentioned with their phylogenetic groups denominated as Cluster I, II and III.

## 4. Discussion

Rapid development of antibiotic-resistance in Gram-negative bacteria including *P. aeruginosa* has become a major public health concern all over the world (2, 3). There is an alarming increase in MDR, XDR, and PDR isolates of *P. aeruginosa* (22, 23). In this study, we also observed frequent occurances of MDR among the *P. aeruginosa* isolates in Bangladesh. Remarkably, one in every three isolates was resistant to carbapenems. While the interquartile range (IQR) of imipenem resistance found in a systematic review covering studies during 2004 to 2018 was 13.5%, we detected a huge increase to 30%(6).

Our results indicate that the rapid increase in imipenem resistance among *P. aeruginosa* clinical isolates in Bangladesh is mediated by MBLs. About 88% of the resistant isolates were positive for MBL production. The MBL-positive isolates also showed an elevated level of MIC values of imipenem and meropenem. Resistance to carbapenems is often associated with the acquisition and expression of the MBLs, which includes *bla*-IMP, *bla*-VIM, *bla-*GIM, *bla*-SPM, *bla*-NDM and *bla*-SIM genes (24). In this study 71% of the carbapenem-resistant isolates were found to carry variants of MBL genes. While the majority (7/16; 44%) of the isolates carried *bla*-VIM, only 19% (3/16) carried *bla-*NDM-1. The current investigation shows a much higher prevalence of MBL genes among *P. aeruginosa* isolates compared to similar studies conducted in India (12%), Pakistan (18%), China (55%), Iran (40%), and Nigeria (17%)(25–28). Moreover, it needs to be noted that we investigated the presence of only four variants (*bla*-IMP, *bla*-VIM, *bla*-SPM, and *bla*-NDM-1). Therefore, we cannot exclude the presence of other MBL variants in remaining 29% of the isolates.

In this study, *bla*-VIM variant was found to be the most prevalent among the MBL genes tested (7/16, 44%). Dissemination of novel variants of *bla-*VIM is commonly mediated by class I integrons like *In58* and *In59* gene cassettes (29, 30). Plasmids carrying class I integrons possess strong dissemination potential (31, 32). However, several of the *bla-*VIM carrier isolates (3/7, 42%) in our study did not carry any plasmid, implying that these isolates may have integrons in their chromosomal DNA.

In this study, the presence of *bla-*NDM-1 was detected among 3 of 16 carbapenem-resistant isolates. Classically *bla-*NDM-1 encoding gene is also plasmid-borne, which has been found to be disseminated through the members of the *Enterobacteriacea*e and *Pseudomonaceae* family (32, 33). Transmission of *bla-*NDM-1 has been also reported in *Acinetobacter baumanii* by transposon Tn125 (34, 35). It has been shown that *bla-*NDM-1 gene can be carried on plasmids of diverse sizes with incompatibility types that allow interspecies, intergenus, and interfamily transfer (36). In this study, we observed the presence of a 0.5 MDa plasmid in *P. aeruginosa* isolates that carried the *bla-*NDM-1 gene. Remarkably, all *bla-*NDM-1 carriers were isolated from UTI and catheter-associated infections.

We employed ERIC-PCR for molecular typing of genetic relatedness among the *P. aeruginosa* isolates. ERIC-PCR revealed that the carbapenem-resistant *P. aeruginosa* isolates were distributed in three distinct clusters, with the majority belonging to cluster III. Overall analyses of our ERIC-PCR data indicated a heterogeneous nature of *P. aeruginosa* isolates and possible independent sources of dissemination. Several previous studies also reported similar heterogenous nature of *P. aeruginosa* isolates (26, 37). However, the XDR isolates found in this study were all grouped in cluster III in ERIC-PCR analyses.

Our test samples were collected from two academic hospitals. Interestingly, both carbapenem resistance and the presence of MBL pattern was different between the two hospitals. Although 35% (6/17) of the *P. aeruginosa* from GMCH were carbapenem resistant, 2 were phenotypically MBL negative and none were positive for the presence of MBL variants tested. In contrast, 28% (10/36) of the *P. aeruginosa* from EMCH were carbapenem resistant, but all the isolates carried MBL genes. Compared to GMCH, EMCH is a larger, tertiary care hospital that admits more patients for a longer period, which might establish a favorable niche for dissemination of MBL genes among nosocomial agents like *P. aeruginosa*. Previous studies also suggested that larger number of patients may harbor frequent source of drug resistant bacterial infections and prolonged hospitalization may increase the risk of acquisition of drug resistance elements through horizontal gene transfer (38, 39). A recent study in Bangladesh also demonstrated that hospital environment is a major reservoir for carbapenem-resistant *P. aeruginosa* with MBL carriage (40).

Indiscriminate use of therapeutically important antibiotics is presumed to be the major cause of antibiotic resistance and the transmission of such resistant pathogens is more likely among hospitalized patients, especially those who need prolonged stay. Horizontal transfer of antibiotic resistance determinants further complicates the control strategies. Our results suggest that rapid screening by molecular techniques like ERIC-PCR may help track multidrug-resistant *P. aeruginosa* outbreaks in hospitals and develop point-of-care surveillance.

## 5. Conclusions

The emerging threat of ever-increasing antibiotic resistance among *P. aeruginosa* of clinical origin is a major concern for the treatment of nosocomial infections caused by the pathogen. The magnitude of multidrug resistance and the presence of MBL variants in clinical settings reported in this study require immediate attention to improve infection control and prevention programs. This study also suspects the hospital environment to facilitate the horizontal transfer of resistance elements like MBL genes. Implementation of antibiotic stewardship policy alongside proper infection control now has become a public health emergency to combat the emerging threat of superbugs in healthcare settings of developing countries like Bangladesh.

## 6. Ethics statement

This study was approved by the Ethics and Research Review Committee of the Jahangirnagar University Faculty of Biological Sciences [Ref No: BBEC, JU/M 2020 (1)4]. Written informed consent was obtained from patients for sample collection, and their personal identities along with other information were anonymized.

## Author Contributions

All authors made a significant contribution to the work reported. M.H.R. conceptualized and supervised the study. H.A., S.Y.M., M.S.A, and M.J.M. collected the clinical samples and conducted laboratory experiments. H.A. and S.Y.M. also analyzed the data and prepared the manuscript. M.S.H. and S.I. made significant contributions to the study design and M.A.K.R. edited the manuscript. The authors agree and approve the content of the final version of the manuscript.

## Data Availability

All data produced in the present study are available upon reasonable request to the authors

## Acknowledgments

The authors would like to thank the laboratory personnel of Enam Medical College Hospital and Gonoshasthaya Medical College Hospital, Dhaka, Bangladesh, for their support in the collection of the clinical samples.

